# Technology acceptance of machine learning in life sciences: the role of hype perception and journal impact factor

**DOI:** 10.64898/2026.06.03.26354262

**Authors:** Antonio E. Serrano

## Abstract

Machine learning (ML) has emerged as a transformative technology across biomedical and life science sectors, with applications spanning drug discovery, medical imaging, genomics, and clinical decision support (Goecks et al., 2020; Patel et al., 2020). Despite exponential growth in ML-related publications, from fewer than 100 articles in 2003 to nearly 25,000 by 2021 (NCBI, 2022), adoption among industry professionals remains uneven and sector-dependent. Understanding what drives or inhibits this adoption is critical for organisations seeking to leverage ML capabilities in research and clinical practice.

Technology adoption in organisational contexts has been extensively studied through the Technology Acceptance Model (TAM), originally proposed by Davis (1989) and subsequently extended to incorporate external variables influencing perceived usefulness (PU) and perceived ease of use (PEU) (Venkatesh & Davis, 1996). While TAM has been applied across multiple industries, its application within biomedical and life science contexts remains limited, and the industry-specific factors that shape ML acceptance in this sector have not been systematically examined.

Two external variables are particularly relevant to life science professionals. First, the bibliometric journal impact factor (JIF) functions as a cognitive signal of scientific credibility, a sector where evidence-based decision-making is culturally embedded, and publication quality serves as a proxy for technological legitimacy (Garfield, 1996). Second, technology hype, operationalised through the Gartner Hype Cycle framework, represents a social influence variable that shapes organisational expectations and investment decisions around emerging technologies (Gartner Inc., 2018). Whether these variables influence ML acceptance among life science professionals, alongside individual knowledge and experience, has not been empirically tested.

This study addresses that gap by investigating ML technology acceptance among 213 biomedical and life science professionals across EMEA, LATAM, and North America, using a cross-sectional quantitative survey and PLS-SEM analysis. The TAM model is extended with three external variables, JIF, technology hype, and prior knowledge and experience, to test their influence on PU and PEU in this specific professional context. Additionally, the study examines demographic and regional differences in ML acceptance, with particular attention to variation between academic researchers and healthcare professionals.

The findings contribute a validated, sector-specific extension of TAM for life sciences, provide actionable insights for organisations seeking to accelerate ML implementation, and establish a framework for future subsector-specific research.

## 1. Literature Review

### 1.1 Machine learning in biomedical and life sciences

The integration of ML into biomedical and life science research has accelerated substantially over the past two decades. ML algorithms have demonstrated utility across a broad range of applications, including drug discovery (Patel et al., 2020), medical imaging analysis (Rajkumar, 2020), genomic data interpretation (Huang et al., 2021), biomarker identification (Shi, Lin & Zhao, 2021), and natural language processing of clinical records (Gong, 2018). The transformation of biology into an information science, driven in part by the completion of the human genome project, has created vast datasets amenable to ML-based analysis (Baltimore, 2001; Nurk et al., 2021).

Despite these advances, adoption of ML tools among biomedical professionals remains inconsistent. Clinicians report difficulties integrating ML into diagnostic workflows (Pumplun et al., 2021), and surveys of physicians and medical students indicate positive but reserved attitudes toward clinical AI applications (Chen et al., 2022). The lack of algorithmic transparency has been identified as a significant barrier to adoption in highly regulated environments (Benchaaben et al., 2020). These observations suggest that technical capability alone does not determine adoption, behavioural and perceptual factors play a critical role.

### 1.2 Technology Acceptance Model

The Technology Acceptance Model (TAM), originally developed by Davis (1985, 1989), provides a well-validated theoretical framework for understanding user adoption of information technologies. The core model posits that two primary perceptions, perceived usefulness (PU) and perceived ease of use (PEU), determine behavioural intention to use (BIU) a technology, which in turn predicts actual technology use (ATU). The relationship between PEU and PU is also established, with ease of use influencing usefulness perceptions.

The extended TAM (Venkatesh & Davis, 1996) introduced external variables as antecedents to PU and PEU, recognising that cognitive and social factors beyond the technology itself shape user perceptions. This extension has been applied across diverse technological contexts, including mobile applications (Alalwan et al., 2018), internet banking (Kaur & Malik, 2019), and e-learning platforms (Jimenez et al., 2021). TAM has also been applied to AI-related technologies, with perceived usefulness and ease of use consistently emerging as significant predictors of adoption intention (Zhang, Wang & Li, 2021).

However, TAM applications within the biomedical and life science industry remain scarce, and the specific cognitive and social variables relevant to this professional context have not been systematically incorporated into the model. Life science professionals operate within a distinctive epistemic culture, one shaped by scientific publishing norms, technology hype cycles, and evidence-based decision-making, that may introduce unique external influences on technology acceptance not captured by generic TAM extensions.

### 1.3 External variables

#### 1.3.1 Journal impact factor as a cognitive variable

The journal impact factor (JIF) is a bibliometric index widely used in life sciences to evaluate the quality and credibility of scientific publications (Garfield, 1996). Given that approximately 77% of respondents in this study held postgraduate degrees, the JIF represents a familiar cognitive reference point for evaluating the scientific legitimacy of emerging technologies. Research published in high-impact journals typically receives greater institutional attention and shapes technology adoption decisions among scientifically trained professionals (Fiala, Mareš & Šesták, 2017). This study proposes JIF as an industry-specific cognitive external variable influencing PU and PEU in the context of ML adoption.

#### 1.3.2 Technology hype as a social variable

The Gartner Hype Cycle provides a framework for understanding how emerging technologies move through phases of inflated expectations, disillusionment, and eventual productivity (Gartner Inc., 2018). In the life science industry, technology hype functions as a social influence variable, shaping peer expectations, investment decisions, and competitive pressures around ML adoption (Sodhi et al., 2022). By 2020, Gartner reported ML entering the trough of disillusionment in general IT sectors, yet life science applications may follow a different trajectory given sector-specific adoption dynamics (Goasduff, 2020). This study operationalises technology hype perception as a social external variable within the TAM framework.

#### 1.3.3 Knowledge and experience

Prior knowledge and experience with a technology have been identified as significant antecedents of PU and PEU across multiple TAM studies (Horowitz & Kahn, 2021). Familiarity with ML concepts, including the ability to distinguish ML from adjacent fields such as deep learning and artificial intelligence, may reduce perceived complexity and increase confidence in technology adoption. This variable is particularly relevant in life sciences, where AI terminology is frequently misused, creating potential misconceptions that influence adoption behaviour (Vincent, 2019).

### 1.4 Hypotheses

Based on the extended TAM framework (Venkatesh & Davis, 1996) and the external variables identified above, ten hypotheses were developed. The core TAM hypotheses (H1–H4) follow established relationships between PEU, PU, BIU, and ATU. Hypotheses H5–H10 test the influence of technology hype (H5, H6), journal impact factor (H7, H8), and knowledge and experience (H9, H10) on PU and PEU respectively. All hypotheses are directional and predict positive relationships between constructs.

## 2. Methods

### 2.1 Research design

This study adopted a cross-sectional quantitative monomethod design, consistent with a positivist epistemological position and deductive research approach (Saunders, Lewis & Thornhill, 2009). A self-administered online survey was used to collect primary data from biomedical and life science professionals. This design is appropriate for testing theoretically derived hypotheses across a geographically dispersed professional population where experimental manipulation is not feasible.

### 2.2 Population and sampling

The target population comprised biomedical and life science professionals, including academic researchers, applied biotechnology scientists, clinical professionals, and pharmaceutical industry practitioners. Convenience sampling was employed, with the survey distributed via the corresponding author’s LinkedIn professional network and direct email contacts. From a total reachable population of 1,347 professionals, 213 completed responses were obtained, representing a response rate of 15.8%. Respondents were drawn primarily from EMEA (57%) and LATAM (30%), with additional representation from North America (10%) and APAC (2%). The two most represented countries were the United Kingdom (34%) and Chile (29%). The sample was predominantly postgraduate-educated (77%), with doctoral degree holders comprising 57% of respondents. The majority worked in the private sector (65%), with academic and research institutions representing 25%.

### 2.3 Survey instrument

The survey instrument was developed using the TAM (Venkatesh & Davis, 1996) as the baseline model and structured into three sections. The first section collected demographic information, including age, gender, country of residence, educational level, industry sector, institution type, and seniority level. The second section measured the core TAM constructs, perceived usefulness (PU, 6 items), perceived ease of use (PEU, 6 items), behavioural intention to use (BIU, 4 items), and actual technology use (ATU, 2 items), using a five-point Likert scale ranging from strongly disagree to strongly agree. The third section assessed the three external variables: journal impact factor (JIF, 3 items), technology hype (TH, 4 items), and knowledge and experience (KE, 3 items), also measured on a five-point Likert scale. An additional item assessed respondents’ perception of ML maturity using a Gartner Hype Cycle-aligned classification. The survey was administered bilingually in English and Spanish to facilitate participation across EMEA and LATAM regions. In total, the instrument comprised 28 Likert-scale items plus demographic questions.

### 2.4 Ethics

Data collection took place between December 20, 2022, and January 15, 2023. Ethics approval was granted by the University of Winchester Research Ethics Committee on December 20, 2022 (Research Ethics Form 3, supervisor: T. Friesner, Faculty of Business and Digital Technologies).

Informed consent was obtained from all participants via an introductory consent statement presented at the beginning of the online survey before data collection. Participation was entirely voluntary and anonymous. No personally identifiable information was collected. No minors were included in the study.

### 2.5 Data analysis

Data analysis was conducted in two stages. First, descriptive statistics and frequency analysis of demographic variables were performed using Microsoft Excel and SPSS Version 27. Second, to test the measurement model and structural hypotheses, Partial Least Squares Structural Equation Modelling (PLS-SEM) was applied using SmartPLS 4 (Ringle, Wende & Becker, 2022). PLS-SEM was selected over covariance-based SEM given the exploratory nature of the extended model, the inclusion of formative constructs, and the relatively modest sample size (Hair et al., 2019). Data were treated as ordinal, and the standard PLS-SEM algorithm was applied.

Measurement model assessment followed established criteria: internal consistency was evaluated using Cronbach’s alpha and composite reliability, with thresholds of 0.7 and above considered acceptable. Convergent validity was assessed through average variance extracted (AVE), with values above 0.5 indicating adequate validity. Discriminant validity was established using cross-loading analysis and the Fornell-Larcker criterion (Fornell & Larcker, 1981). Structural model assessment reported standardised path coefficients (β), R-squared values, predictive relevance (Q²), and effect sizes (f²). Hypothesis testing was based on t-statistics and p-values, with p ≤ 0.05 adopted as the significance threshold.

To further explore demographic influences on TAM constructs, Principal Component Analysis (PCA), Exploratory Factor Analysis (EFA) with Varimax rotation, Multiple Correspondence Analysis (MCA), and t-SNE were performed using the FactoMineR package in R (version 4.2.2) and SPSS Version 27.

## 3. Results

### 3.1 Respondent characteristics

The sample comprised 213 biomedical and life science professionals. Gender distribution was approximately balanced, with male respondents representing 52% and female 47%. The majority of respondents held doctoral degrees (57%), followed by master’s degrees (20%) and professional qualifications (10%). In terms of industry sector, applied biotechnology represented the largest group (47%), followed by basic biological research (16%), healthcare diagnostics (14%), and pharmaceuticals (12%). Most respondents worked in private sector organisations (65%), with academic and research institutions accounting for 25%. Regionally, EMEA represented 57% of respondents, LATAM 30%, North America 10%, and APAC 2%. The 35–44 age bracket was the most represented (41%), followed by 45–54 (32%). Full respondent characteristics are presented in Table 1.

**Table 1:** Respondent Characteristics.

|  | Category | Frequency | Percentage |
| --- | --- | --- | --- |
| **Preferred Language** | English | 121 | 58% |
|  | Spanish | 89 | 42% |
| **Gender** | Female | 99 | 47% |
|  | Male | 109 | 52% |
|  | Prefer not to say | 2 | 1% |
| **Educational Level** | School qualification | 3 | 1% |
|  | Bachelor degree | 27 | 13% |
|  | Doctorate | 119 | 57% |
|  | Master degree | 41 | 20% |
|  | Professional qualification | 20 | 10% |
| **Industry Area** | Applied biotechnology - nonclinical | 56 | 27% |
|  | Applied biotechnology in health and biomedicine | 43 | 20% |
|  | Basic biological research | 33 | 16% |
|  | Healthcare - Point of care - Diagnostics | 30 | 14% |
|  | Pharmaceuticals. Development or manufacturing | 26 | 12% |
|  | Other | 22 | 10% |
| **Organisation Type** | Company - For-profit organisation - Commercial entity | 136 | 65% |
|  | Public or private education/research - University | 52 | 25% |
|  | Hospital or Clinical institution | 18 | 9% |
|  | Other | 4 | 2% |
| **Region** | APAC | 5 | 2% |
|  | EMEA | 120 | 57% |
|  | LATAM | 64 | 30% |
|  | NAM | 21 | 10% |
| **Age** | 18 - 24 | 2 | 1% |
|  | 25 - 34 | 34 | 16% |
|  | 35 - 44 | 87 | 41% |
|  | 45 - 54 | 68 | 32% |
|  | 55 - 64 | 17 | 8% |
|  | 65 or over | 2 | 1% |

### 3.2 Measurement model

Reliability and validity of the measurement model were assessed prior to hypothesis testing. All constructs exceeded the recommended Cronbach’s alpha threshold of 0.7, with values ranging from 0.799 (TH) to 0.939 (PU). Composite reliability values similarly exceeded 0.85 for all constructs. Average variance extracted (AVE) values exceeded the 0.5 threshold for all constructs, ranging from 0.630 (TH) to 0.855 (ATU), confirming convergent validity (Table 2).

**Table 2:**
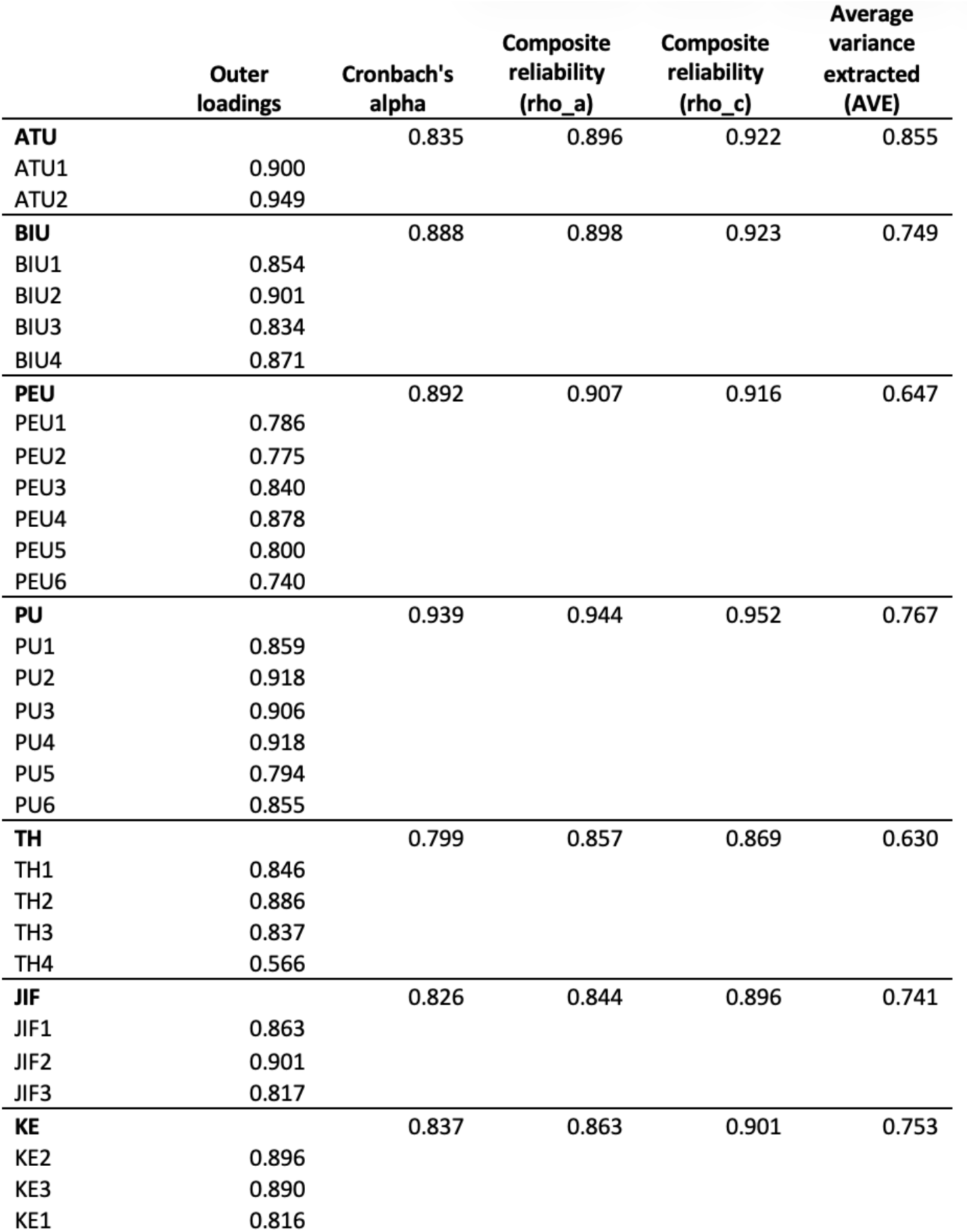
Validity and Reliability.

Discriminant validity was established through cross-loading analysis, which confirmed that all indicator loadings were higher on their respective constructs than on any other construct (Table 3). The Fornell-Larcker criterion was also satisfied, with the square root of each construct’s AVE exceeding its correlations with all other constructs (Table 4). The ATU construct showed the highest discriminant validity (0.925). These results confirm that the measurement model meets established standards of reliability and validity.

**Table 3.** Cross-loading.

|  | ATU | BIU | JIF | PEU | PU | TH | KE |
| --- | --- | --- | --- | --- | --- | --- | --- |
| ATU1 | 0.900 | 0.338 | 0.233 | 0.323 | 0.339 | 0.477 | 0.333 |
| ATU2 | 0.949 | 0.465 | 0.337 | 0.391 | 0.462 | 0.410 | 0.441 |
| BIU1 | 0.334 | 0.854 | 0.235 | 0.493 | 0.554 | 0.286 | 0.340 |
| BIU2 | 0.465 | 0.901 | 0.317 | 0.521 | 0.643 | 0.353 | 0.385 |
| BIU3 | 0.300 | 0.834 | 0.240 | 0.484 | 0.493 | 0.252 | 0.261 |
| BIU4 | 0.417 | 0.871 | 0.302 | 0.428 | 0.541 | 0.390 | 0.343 |
| PEU1 | 0.310 | 0.548 | 0.246 | 0.786 | 0.370 | 0.263 | 0.295 |
| PEU2 | 0.372 | 0.390 | 0.250 | 0.775 | 0.276 | 0.226 | 0.251 |
| PEU3 | 0.294 | 0.479 | 0.189 | 0.840 | 0.415 | 0.244 | 0.203 |
| PEU4 | 0.374 | 0.485 | 0.214 | 0.878 | 0.441 | 0.327 | 0.224 |
| PEU5 | 0.227 | 0.434 | 0.115 | 0.800 | 0.266 | 0.145 | 0.216 |
| PEU6 | 0.297 | 0.270 | 0.137 | 0.740 | 0.199 | 0.144 | 0.218 |
| PU1 | 0.348 | 0.563 | 0.227 | 0.357 | 0.859 | 0.325 | 0.265 |
| PU2 | 0.402 | 0.582 | 0.275 | 0.404 | 0.918 | 0.282 | 0.294 |
| PU3 | 0.356 | 0.571 | 0.351 | 0.342 | 0.906 | 0.337 | 0.210 |
| PU4 | 0.403 | 0.558 | 0.270 | 0.376 | 0.918 | 0.327 | 0.313 |
| PU5 | 0.392 | 0.463 | 0.282 | 0.339 | 0.794 | 0.276 | 0.231 |
| PU6 | 0.418 | 0.645 | 0.335 | 0.404 | 0.855 | 0.413 | 0.392 |
| TH1 | 0.360 | 0.247 | 0.150 | 0.253 | 0.270 | 0.846 | 0.188 |
| TH2 | 0.425 | 0.355 | 0.268 | 0.281 | 0.394 | 0.886 | 0.230 |
| TH3 | 0.394 | 0.319 | 0.279 | 0.247 | 0.285 | 0.837 | 0.239 |
| TH4 | 0.319 | 0.256 | 0.249 | 0.101 | 0.214 | 0.566 | 0.224 |
| JIF1 | 0.191 | 0.258 | 0.863 | 0.197 | 0.248 | 0.243 | 0.113 |
| JIF2 | 0.306 | 0.341 | 0.901 | 0.218 | 0.349 | 0.313 | 0.115 |
| JIF3 | 0.309 | 0.209 | 0.817 | 0.219 | 0.249 | 0.188 | 0.093 |
| KE1 | 0.251 | 0.323 | 0.105 | 0.259 | 0.213 | 0.219 | 0.816 |
| KE2 | 0.525 | 0.391 | 0.113 | 0.280 | 0.354 | 0.266 | 0.896 |
| KE3 | 0.285 | 0.282 | 0.104 | 0.217 | 0.270 | 0.214 | 0.890 |

**Table 4:** Fornell-Larcker Criterion.

|  | ATU | BIU | PEU | PU | TH | JIF | KE |
| --- | --- | --- | --- | --- | --- | --- | --- |
| ATU | **0.925** |  |  |  |  |  |  |
| BIU | 0.443 | **0.865** |  |  |  |  |  |
| PEU | 0.390 | 0.557 | **0.804** |  |  |  |  |
| PU | 0.442 | 0.649 | 0.425 | **0.876** |  |  |  |
| TH | 0.472 | 0.373 | 0.291 | 0.377 | **0.794** |  |  |
| JIF | 0.316 | 0.318 | 0.245 | 0.333 | 0.293 | **0.861** |  |
| KE | 0.426 | 0.388 | 0.293 | 0.329 | 0.271 | 0.124 | **0.868** |

### 3.3 Structural model

The structural model was assessed through R-squared values, predictive relevance (Q²), and effect sizes (f²) (Table 5). The BIU construct demonstrated the highest R-squared value (0.517), indicating moderate explanatory power. PU showed a weak R-squared (0.308), while ATU (0.197) and PEU (0.159) showed insufficient predictive acceptance as outcome variables. Q² values exceeded zero for all constructs, BIU (0.236), PU (0.218), PEU (0.122), confirming moderate predictive relevance across the model. The largest effect size was observed for PU on BIU (f² = 0.429), consistent with established TAM findings (Table 6).

**Table 5:** Outcome variables and Predictive Relevance Q^2^.

| | R-square | R-square adjusted | Criteria | $Q^2$predict | Criteria |
| --- | --- | --- | --- | --- | --- |
| ATU | 0.197 | 0.193 | Insufficient | 0.166 | Moderate |
| BIU | 0.517 | 0.513 | Moderate | 0.236 | Moderate |
| PEU | 0.159 | 0.147 | Insufficient | 0.122 | Moderate |
| PU | 0.308 | 0.295 | Weak | 0.218 | Moderate |

**Table 6:**
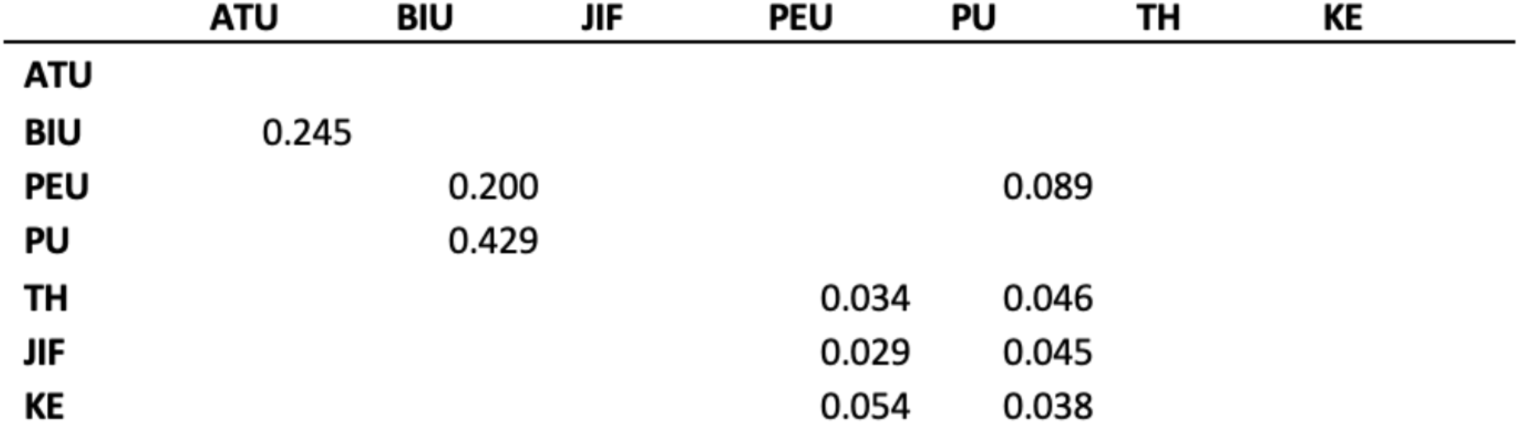
Effect Size f2.

|  | ATU | BIU | JIF | PEU | PU | TH | KE |
| --- | --- | --- | --- | --- | --- | --- | --- |
| ATU |  |  |  |  |  |  |  |
| BIU | 0.245 |  |  |  |  |  |  |
| PEU |  | 0.200 |  |  | 0.089 |  |  |
| PU |  | 0.429 |  |  |  |  |  |
| TH |  |  |  | 0.034 | 0.046 |  |  |
| JIF |  |  |  | 0.029 | 0.045 |  |  |
| KE |  |  |  | 0.054 | 0.038 |  |  |

### 3.4 Hypothesis testing

All ten hypotheses were supported at p ≤ 0.05 (Table 7). The strongest path coefficient was observed for PU → BIU (β = 0.503, p < 0.001), confirming perceived usefulness as the primary driver of behavioural intention to use ML. PEU → PU (β = 0.271, p < 0.001) and PEU → BIU (β = 0.344, p < 0.001) were also significant, establishing the core TAM structure. BIU → ATU (β = 0.443, p < 0.001) confirmed the relationship between intention and actual use.

**Table 7:** Hypothesis Testing.

| Hypothesis | Path | Path coefficients | Original sample (O) | Sample mean (M) | Standard deviation (STDEV) | T statistics (\|O/STDEV\|) | P values | Decision |
| --- | --- | --- | --- | --- | --- | --- | --- | --- |
| H1 | PEU -> PU | 0.271 | 0.271 | 0.269 | 0.063 | 4.282 | 0.000 | Accepted |
| H2 | PU -> BIU | 0.503 | 0.503 | 0.505 | 0.055 | 9.155 | 0.000 | Accepted |
| H3 | PEU -> BIU | 0.344 | 0.344 | 0.344 | 0.052 | 6.619 | 0.000 | Accepted |
| H4 | BIU -> ATU | 0.443 | 0.443 | 0.445 | 0.050 | 8.900 | 0.000 | Accepted |
| H5 | TH -> PU | 0.196 | 0.196 | 0.197 | 0.067 | 2.929 | 0.003 | Accepted |
| H6 | TH -> PEU | 0.183 | 0.183 | 0.182 | 0.070 | 2.599 | 0.009 | Accepted |
| H7 | JIF -> PU | 0.187 | 0.187 | 0.191 | 0.065 | 2.883 | 0.004 | Accepted |
| H8 | JIF -> PEU | 0.164 | 0.164 | 0.168 | 0.076 | 2.156 | 0.031 | Accepted |
| H9 | KE -> PU | 0.174 | 0.174 | 0.175 | 0.055 | 3.158 | 0.002 | Accepted |
| H10 | KE -> PEU | 0.223 | 0.223 | 0.226 | 0.069 | 3.212 | 0.001 | Accepted |

Regarding external variables, technology hype positively influenced both PU (β = 0.196, p = 0.003) and PEU (β = 0.183, p = 0.009), supporting H5 and H6. Journal impact factor showed significant positive effects on PU (β = 0.187, p = 0.004) and PEU (β = 0.164, p = 0.031), supporting H7 and H8. Knowledge and experience positively influenced PU (β = 0.174, p = 0.002) and PEU (β = 0.223, p = 0.001), supporting H9 and H10.

### 3.5 Demographic and factor analyses

Principal Component Analysis revealed that language and GNI per capita were the variables with the highest contribution to variance differences among respondents, while age, gender, education, and seniority showed minimal contribution (Figure 2). No significant regional clustering was identified between LATAM and North Atlantic respondents in ML acceptance, suggesting a homogeneous adoption profile across the western hemisphere among highly educated professionals.

**Figure 1.**
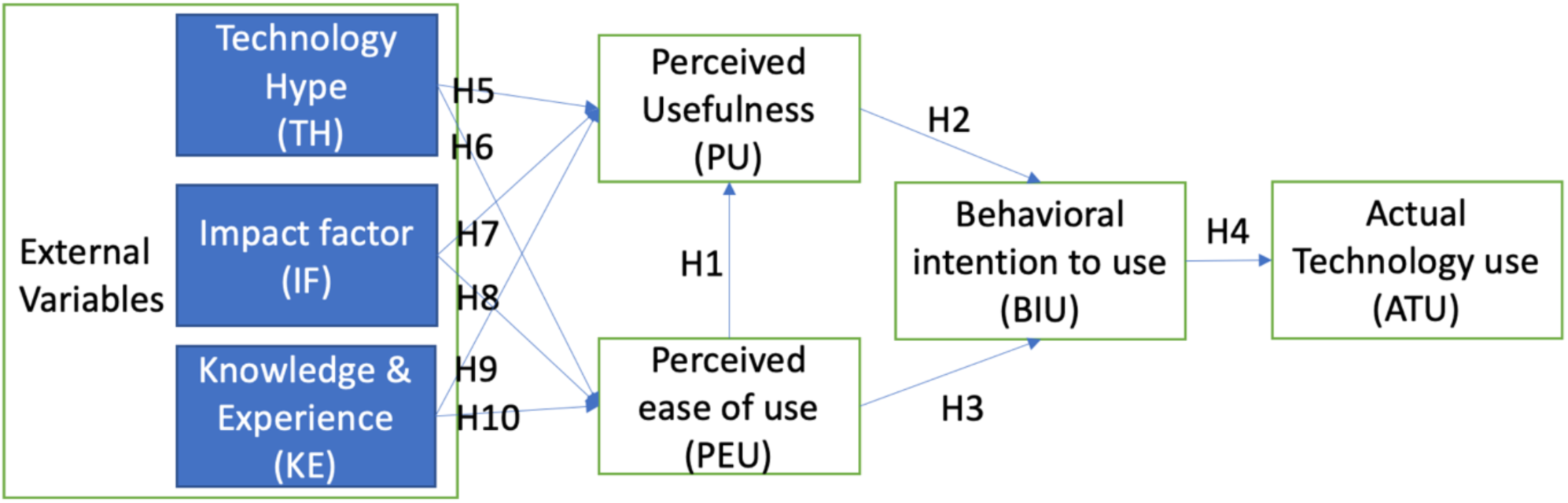
The theoretical framework of the research study. Three external factors are proposed to integrate as external variables for adopting ML technologies in the life sciences industry: the scientometric perception of research impact, the general industry perception from a hype cycle point of view, and knowledge and previous experience in the field (Venkatesh and Davis, 1996)

**Figure 2.**
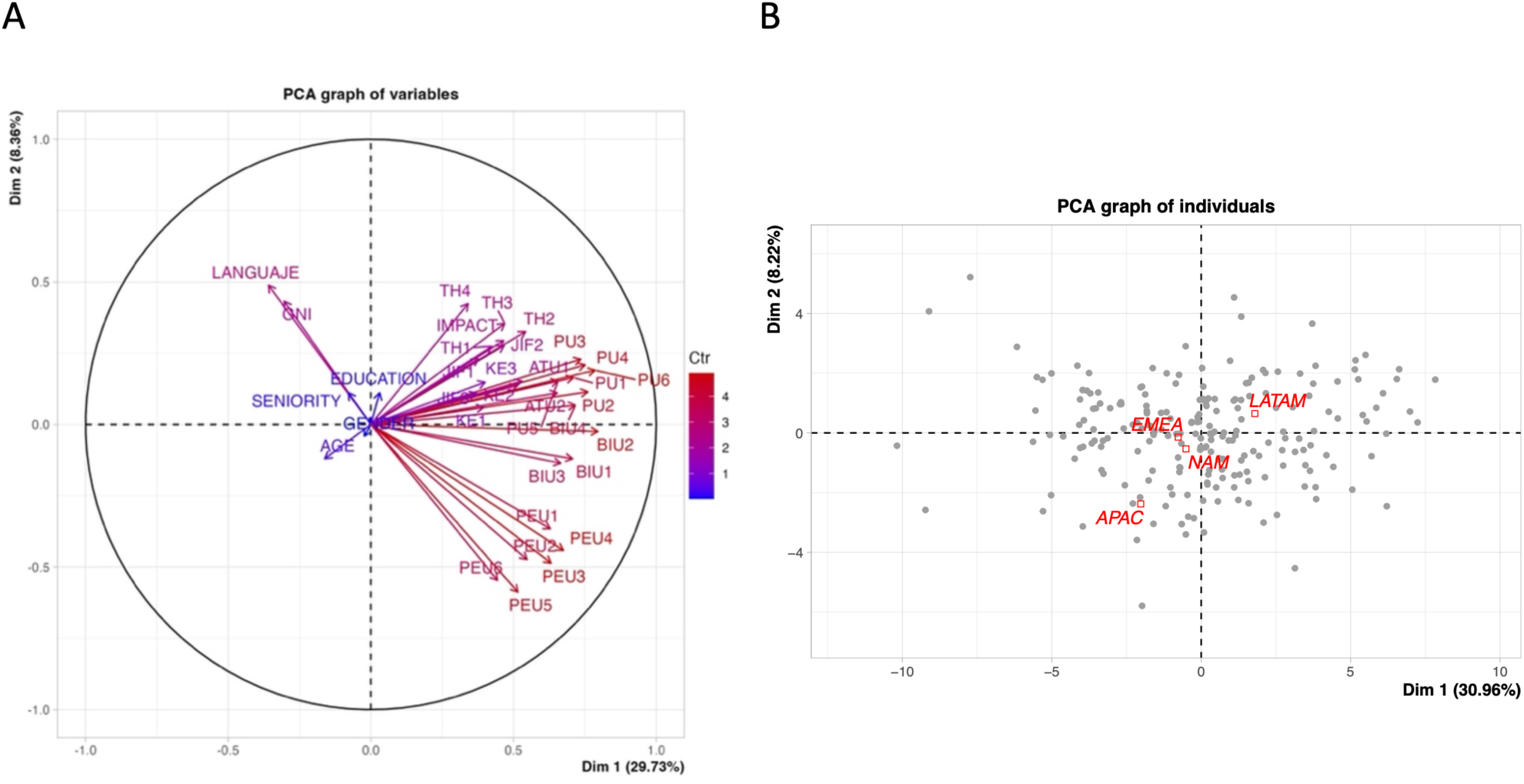
Principal Component Analysis: A) Variables with the highest contribution to the variance differences are language and GNI. Education, seniority level, gender and age are the lowest contributors. B) PCA graph of individuals, compared with the Region qualitative variable. The total percentage of explained components was 39.18-38.09%.

Exploratory Factor Analysis identified eight underlying components explaining 62.8% of total variance, with the first component, comprising ATU, BIU, and PU items, accounting for 30% of variance (Table 8). Demographic variables were insufficient to explain differences in TAM construct scores, confirming that individual professional characteristics do not significantly moderate ML acceptance in this sample.

**Table 8:** Heat Map of the Varimax bivariate Spearman two-tailed non-parametric correlation.

|  | GNI | Language | Age | Gender | Education | Seniority | ATU1 | ATU2 | BIU1 | BIU2 | BIU3 | BIU4 | PEU1 | PEU2 | PEU3 | PEU4 | PEU5 | PEU6 | PU1 | PU2 | PU3 | PU4 | PU5 | PU6 | TH1 | TH2 | TH3 | TH4 | JIF1 | JIF2 | JIF3 | KE1 | KE2 | KE3 | Impact |
| --- | --- | --- | --- | --- | --- | --- | --- | --- | --- | --- | --- | --- | --- | --- | --- | --- | --- | --- | --- | --- | --- | --- | --- | --- | --- | --- | --- | --- | --- | --- | --- | --- | --- | --- | --- |
| GNI | 1.0 |  |  |  |  |  |  |  |  |  |  |  |  |  |  |  |  |  |  |  |  |  |  |  |  |  |  |  |  |  |  |  |  |  |  |
| Language | 0.7 | 1.0 |  |  |  |  |  |  |  |  |  |  |  |  |  |  |  |  |  |  |  |  |  |  |  |  |  |  |  |  |  |  |  |  |  |
| Age | -0.1 | -0.1 | 1.0 |  |  |  |  |  |  |  |  |  |  |  |  |  |  |  |  |  |  |  |  |  |  |  |  |  |  |  |  |  |  |  |  |
| Gender | 0.0 | 0.0 | 0.1 | 1.0 |  |  |  |  |  |  |  |  |  |  |  |  |  |  |  |  |  |  |  |  |  |  |  |  |  |  |  |  |  |  |  |
| Education | 0.0 | -0.1 | 0.1 | 0.0 | 1.0 |  |  |  |  |  |  |  |  |  |  |  |  |  |  |  |  |  |  |  |  |  |  |  |  |  |  |  |  |  |  |
| Seniority | 0.0 | 0.1 | 0.3 | 0.2 | 0.1 | 1.0 |  |  |  |  |  |  |  |  |  |  |  |  |  |  |  |  |  |  |  |  |  |  |  |  |  |  |  |  |  |
| ATU1 | -0.1 | -0.1 | 0.0 | -0.1 | 0.1 | -0.1 | 1.0 |  |  |  |  |  |  |  |  |  |  |  |  |  |  |  |  |  |  |  |  |  |  |  |  |  |  |  |  |
| ATU2 | -0.2 | -0.3 | -0.1 | -0.1 | 0.0 | 0.0 | 0.7 | 1.0 |  |  |  |  |  |  |  |  |  |  |  |  |  |  |  |  |  |  |  |  |  |  |  |  |  |  |  |
| BIU1 | -0.3 | -0.3 | -0.1 | 0.0 | 0.1 | 0.0 | 0.2 | 0.4 | 1.0 |  |  |  |  |  |  |  |  |  |  |  |  |  |  |  |  |  |  |  |  |  |  |  |  |  |  |
| BIU2 | -0.3 | -0.3 | -0.1 | 0.0 | 0.1 | 0.0 | 0.4 | 0.5 | 0.7 | 1.0 |  |  |  |  |  |  |  |  |  |  |  |  |  |  |  |  |  |  |  |  |  |  |  |  |  |
| BIU3 | -0.3 | -0.3 | -0.2 | -0.1 | 0.1 | -0.1 | 0.2 | 0.3 | 0.7 | 0.6 | 1.0 |  |  |  |  |  |  |  |  |  |  |  |  |  |  |  |  |  |  |  |  |  |  |  |  |
| BIU4 | -0.2 | -0.2 | -0.2 | 0.0 | 0.0 | 0.0 | 0.3 | 0.4 | 0.6 | 0.7 | 0.6 | 1.0 |  |  |  |  |  |  |  |  |  |  |  |  |  |  |  |  |  |  |  |  |  |  |  |
| PEU1 | -0.2 | -0.3 | -0.1 | 0.1 | 0.1 | 0.0 | 0.2 | 0.3 | 0.5 | 0.5 | 0.4 | 0.5 | 1.0 |  |  |  |  |  |  |  |  |  |  |  |  |  |  |  |  |  |  |  |  |  |  |
| PEU2 | -0.2 | -0.3 | 0.1 | 0.1 | -0.1 | 0.0 | 0.3 | 0.3 | 0.3 | 0.4 | 0.3 | 0.3 | 0.5 | 1.0 |  |  |  |  |  |  |  |  |  |  |  |  |  |  |  |  |  |  |  |  |  |
| PEU3 | -0.3 | -0.4 | 0.0 | 0.0 | -0.1 | -0.1 | 0.2 | 0.3 | 0.5 | 0.4 | 0.4 | 0.4 | 0.5 | 0.5 | 1.0 |  |  |  |  |  |  |  |  |  |  |  |  |  |  |  |  |  |  |  |  |
| PEU4 | -0.3 | -0.3 | -0.1 | 0.0 | 0.0 | -0.1 | 0.3 | 0.4 | 0.4 | 0.5 | 0.4 | 0.3 | 0.6 | 0.5 | 0.7 | 1.0 |  |  |  |  |  |  |  |  |  |  |  |  |  |  |  |  |  |  |  |
| PEU5 | -0.2 | -0.3 | 0.0 | 0.0 | -0.1 | -0.2 | 0.2 | 0.2 | 0.4 | 0.4 | 0.4 | 0.3 | 0.5 | 0.5 | 0.6 | 0.6 | 1.0 |  |  |  |  |  |  |  |  |  |  |  |  |  |  |  |  |  |  |
| PEU6 | -0.2 | -0.2 | 0.0 | 0.0 | -0.2 | 0.0 | 0.3 | 0.2 | 0.3 | 0.2 | 0.2 | 0.1 | 0.4 | 0.5 | 0.5 | 0.5 | 0.6 | 1.0 |  |  |  |  |  |  |  |  |  |  |  |  |  |  |  |  |  |
| PU1 | -0.2 | -0.2 | -0.1 | -0.1 | 0.1 | 0.0 | 0.3 | 0.4 | 0.5 | 0.6 | 0.5 | 0.5 | 0.3 | 0.2 | 0.3 | 0.4 | 0.3 | 0.1 | 1.0 |  |  |  |  |  |  |  |  |  |  |  |  |  |  |  |  |
| PU2 | -0.2 | -0.3 | -0.1 | -0.1 | 0.0 | 0.0 | 0.3 | 0.4 | 0.5 | 0.6 | 0.5 | 0.5 | 0.3 | 0.2 | 0.4 | 0.4 | 0.3 | 0.2 | 0.7 | 1.0 |  |  |  |  |  |  |  |  |  |  |  |  |  |  |  |
| PU3 | -0.1 | -0.2 | -0.1 | 0.0 | 0.0 | 0.0 | 0.2 | 0.4 | 0.5 | 0.6 | 0.5 | 0.5 | 0.3 | 0.1 | 0.4 | 0.4 | 0.2 | 0.2 | 0.7 | 0.8 | 1.0 |  |  |  |  |  |  |  |  |  |  |  |  |  |  |
| PU4 | -0.1 | -0.2 | 0.0 | 0.0 | 0.0 | 0.0 | 0.3 | 0.4 | 0.5 | 0.6 | 0.4 | 0.5 | 0.4 | 0.2 | 0.4 | 0.4 | 0.3 | 0.2 | 0.7 | 0.8 | 0.8 | 1.0 |  |  |  |  |  |  |  |  |  |  |  |  |  |
| PU5 | -0.2 | -0.2 | -0.1 | 0.0 | 0.0 | -0.1 | 0.3 | 0.4 | 0.4 | 0.5 | 0.3 | 0.4 | 0.3 | 0.2 | 0.3 | 0.3 | 0.2 | 0.2 | 0.6 | 0.7 | 0.6 | 0.7 | 1.0 |  |  |  |  |  |  |  |  |  |  |  |  |
| PU6 | -0.2 | -0.2 | -0.1 | 0.0 | 0.1 | -0.1 | 0.3 | 0.4 | 0.6 | 0.6 | 0.6 | 0.6 | 0.3 | 0.3 | 0.4 | 0.4 | 0.3 | 0.2 | 0.7 | 0.7 | 0.7 | 0.7 | 0.6 | 1.0 |  |  |  |  |  |  |  |  |  |  |  |
| TH1 | 0.0 | 0.0 | 0.0 | 0.0 | 0.1 | 0.1 | 0.4 | 0.3 | 0.2 | 0.2 | 0.2 | 0.2 | 0.1 | 0.2 | 0.2 | 0.2 | 0.1 | 0.2 | 0.2 | 0.1 | 0.2 | 0.2 | 0.1 | 0.2 | 1.0 |  |  |  |  |  |  |  |  |  |  |
| TH2 | 0.0 | 0.0 | -0.1 | 0.0 | 0.0 | 0.0 | 0.4 | 0.4 | 0.3 | 0.3 | 0.3 | 0.3 | 0.2 | 0.2 | 0.2 | 0.3 | 0.1 | 0.1 | 0.3 | 0.3 | 0.3 | 0.3 | 0.3 | 0.4 | 0.6 | 1.0 |  |  |  |  |  |  |  |  |  |
| TH3 | 0.0 | 0.1 | -0.1 | 0.1 | 0.1 | 0.0 | 0.4 | 0.4 | 0.2 | 0.3 | 0.2 | 0.3 | 0.2 | 0.2 | 0.2 | 0.2 | 0.1 | 0.1 | 0.2 | 0.2 | 0.3 | 0.2 | 0.1 | 0.3 | 0.6 | 0.6 | 1.0 |  |  |  |  |  |  |  |  |
| TH4 | 0.1 | 0.1 | -0.1 | 0.0 | 0.1 | 0.0 | 0.3 | 0.3 | 0.2 | 0.2 | 0.2 | 0.3 | 0.1 | 0.0 | 0.1 | 0.2 | 0.0 | -0.1 | 0.2 | 0.2 | 0.2 | 0.2 | 0.2 | 0.2 | 0.4 | 0.4 | 0.4 | 1.0 |  |  |  |  |  |  |  |
| JIF1 | 0.0 | 0.0 | -0.1 | 0.0 | 0.0 | -0.1 | 0.1 | 0.2 | 0.2 | 0.3 | 0.2 | 0.3 | 0.2 | 0.2 | 0.1 | 0.1 | 0.1 | 0.0 | 0.2 | 0.2 | 0.3 | 0.2 | 0.2 | 0.3 | 0.1 | 0.2 | 0.2 | 0.2 | 1.0 |  |  |  |  |  |  |
| JIF2 | -0.1 | -0.1 | -0.1 | 0.0 | 0.0 | 0.0 | 0.2 | 0.3 | 0.2 | 0.3 | 0.3 | 0.3 | 0.2 | 0.1 | 0.2 | 0.2 | 0.0 | 0.1 | 0.2 | 0.3 | 0.3 | 0.3 | 0.3 | 0.3 | 0.1 | 0.2 | 0.3 | 0.2 | 0.7 | 1.0 |  |  |  |  |  |
| JIF3 | -0.1 | -0.1 | -0.1 | -0.1 | -0.1 | -0.1 | 0.2 | 0.3 | 0.2 | 0.2 | 0.2 | 0.2 | 0.2 | 0.3 | 0.1 | 0.2 | 0.1 | 0.2 | 0.2 | 0.2 | 0.3 | 0.2 | 0.2 | 0.2 | 0.1 | 0.1 | 0.1 | 0.1 | 0.5 | 0.5 | 1.0 |  |  |  |  |
| KE1 | -0.1 | -0.1 | 0.0 | 0.1 | 0.2 | 0.2 | 0.2 | 0.3 | 0.3 | 0.3 | 0.2 | 0.3 | 0.3 | 0.2 | 0.2 | 0.1 | 0.2 | 0.1 | 0.2 | 0.2 | 0.1 | 0.2 | 0.1 | 0.3 | 0.2 | 0.2 | 0.2 | 0.2 | 0.1 | 0.1 | 0.0 | 1.0 |  |  |  |
| KE2 | 0.0 | 0.0 | -0.1 | 0.0 | 0.1 | 0.1 | 0.4 | 0.5 | 0.3 | 0.4 | 0.3 | 0.4 | 0.3 | 0.2 | 0.2 | 0.2 | 0.2 | 0.2 | 0.3 | 0.4 | 0.2 | 0.4 | 0.3 | 0.4 | 0.1 | 0.3 | 0.2 | 0.3 | 0.1 | 0.1 | 0.1 | 0.5 | 1.0 |  |  |
| KE3 | 0.0 | 0.0 | -0.1 | 0.0 | 0.1 | 0.1 | 0.2 | 0.3 | 0.3 | 0.3 | 0.2 | 0.2 | 0.2 | 0.2 | 0.2 | 0.2 | 0.1 | 0.2 | 0.2 | 0.2 | 0.2 | 0.3 | 0.2 | 0.4 | 0.1 | 0.2 | 0.3 | 0.2 | 0.1 | 0.1 | 0.1 | 0.6 | 0.7 | 1.0 |  |
| Impact | -0.1 | 0.0 | -0.1 | 0.0 | 0.0 | 0.1 | 0.2 | 0.3 | 0.3 | 0.3 | 0.3 | 0.4 | 0.3 | 0.1 | 0.3 | 0.2 | 0.1 | 0.1 | 0.4 | 0.3 | 0.4 | 0.4 | 0.3 | 0.4 | 0.2 | 0.3 | 0.3 | 0.3 | 0.3 | 0.3 | 0.3 | 0.2 | 0.2 | 0.2 | 1.0 |

Multiple Correspondence Analysis revealed a subtle clustering between academic research professionals and healthcare workers, with the latter grouping separately from educational and research organisations (Figure 3). However, the two dimensions explained only 23% of total variance, indicating that the survey instrument did not strongly capture this differentiation.

**Figure 3.**
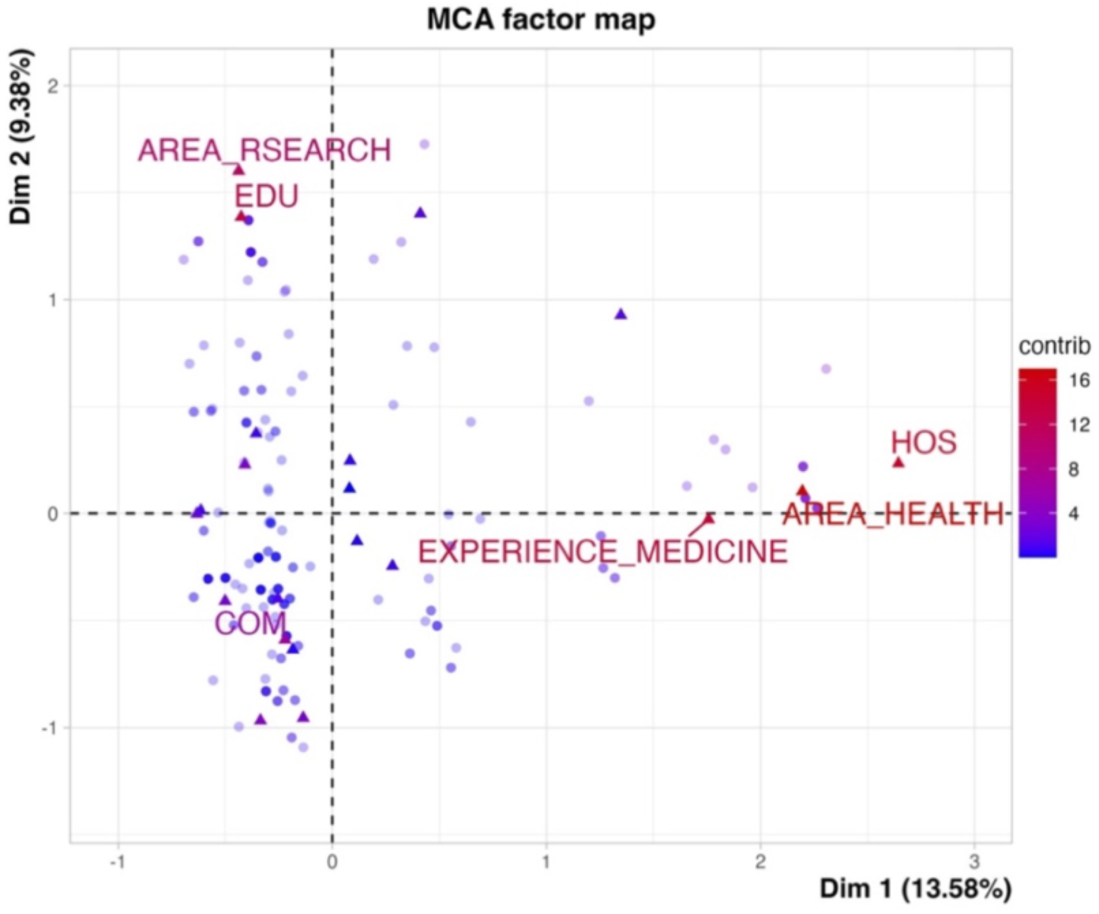
MCA factor map of the individuals surveyed. The qualitative factors were considered: area of occupation in Life Sciences, the field of expertise, and the type of organisation. The highest contributors to the differences are represented. HOS: hospitals, EDU: educational organisations.

### 3.6 Technology hype assessment

Respondents’ perceptions of ML maturity were distributed across the Gartner Hype Cycle stages, with 31% placing ML in early stages, 18% at peak hype, 26% in the growth phase, and only 3% reporting disillusionment (Figure 4). This distribution contrasts with Gartner’s 2020 general IT report, placing ML in the trough of disillusionment, suggesting that ML adoption in biomedical and life science sectors follows a different trajectory than general information technology industries.

**Figure 4:**
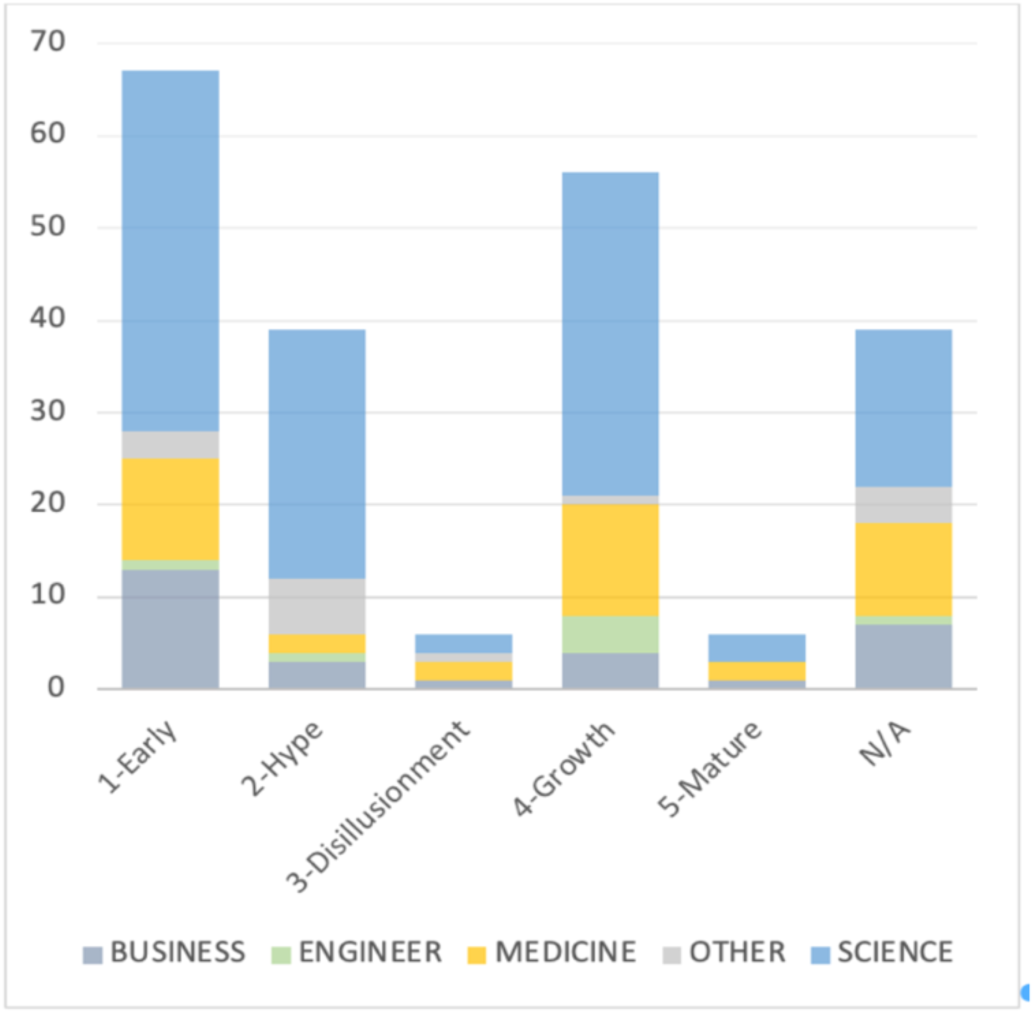
Perception of technology hype of ML among biomedical and health care professionals. Bar plot of respondents by work performance area in the industry.

Healthcare professionals were notably less likely to perceive ML as experiencing hype, and they reported higher technology maturity in their field than academic researchers.

## 4. Discussion

### 4.1 TAM model validation

The results confirm the applicability of the Technology Acceptance Model in the biomedical and life science industry context. All four core TAM constructs, perceived usefulness, perceived ease of use, behavioural intention to use, and actual technology use, were validated, with all hypothesised relationships supported at p ≤ 0.05. These findings are consistent with TAM applications in adjacent technological contexts, including AI adoption (Zhang, Wang & Li, 2021) and mobile health technologies (Alalwan et al., 2018), confirming the robustness of the model across professional and industrial settings.

Behavioural intention to use emerged as the most influential construct in predicting actual ML use (β = 0.443), consistent with established TAM literature (Davis, 1989; Venkatesh & Davis, 1996). This finding has a practical implication, organisations seeking to accelerate ML adoption should prioritise interventions that strengthen professionals’ intention to use, rather than focusing exclusively on technical implementation. Training programmes, peer exposure, and demonstrable use cases that build positive intention are likely to be more effective than technology deployment alone. Perceived usefulness was the strongest predictor of behavioural intention (β = 0.503), reinforcing that life science professionals are pragmatic adopters, they will engage with ML when they perceive a tangible benefit to their work performance and decision-making. This aligns with the sector’s evidence-based culture, where adoption is driven by demonstrated value rather than novelty.

### 4.2 External variables

The acceptance of all three external variable hypotheses represents the primary theoretical contribution of this study. Technology hype, journal impact factor, and knowledge and experience each positively influenced both perceived usefulness and perceived ease of use, confirming their relevance as antecedents of ML acceptance in life science contexts.

The positive influence of technology hype on PU and PEU (H5, H6) suggests that social pressure from peers and competitors plays a meaningful role in shaping ML adoption perceptions among life science professionals. This is consistent with the social influence component of extended TAM models (Venkatesh & Davis, 2000) and with research on observational learning in technology adoption (Gu et al., 2015). Notably, the hype assessment results revealed that life science professionals do not perceive ML as experiencing the disillusionment reported in Gartner’s 2020 general IT report, suggesting that sector-specific hype dynamics differ from broader technology trends, and that social pressure toward ML adoption remains active in this industry.

The positive influence of journal impact factor on PU and PEU (H7, H8) is a novel finding with specific relevance to scientifically trained professionals. The JIF functions as a cognitive proxy for technological legitimacy in a sector where evidence-based decision-making is culturally embedded. Professionals who regularly evaluate research quality through bibliometric signals appear to extend this evaluative framework to technology adoption decisions — perceiving ML as more useful and easier to adopt when its applications are validated in high-impact publications. This finding suggests that disseminating ML research through prestigious scientific journals may accelerate adoption more effectively than industry marketing communications in this specific professional context.

Knowledge and experience showed the strongest influence on PEU (β = 0.223), confirming that prior familiarity with ML reduces perceived complexity and increases confidence in adoption. This is consistent with previous research showing that self-reported AI familiarity correlates with increased approval of AI use (Horowitz & Kahn, 2021). The finding also highlights a critical adoption barrier, professionals who lack prior ML exposure may systematically underestimate its accessibility, suggesting that educational interventions focused on hands-on experience could meaningfully shift adoption perceptions.

### 4.3 Demographic and regional findings

No significant demographic differences in ML acceptance were identified across gender, age, seniority, or region. The absence of regional differences between LATAM and North Atlantic professionals is particularly noteworthy, suggesting that among highly educated life science professionals, ML acceptance converges across geographic and cultural contexts. This contrasts with earlier technology adoption research identifying regional differences (Hultberg et al., 1999), and may reflect the globalised nature of scientific training and publication culture in life sciences.

The homogeneity of the sample, predominantly postgraduate-educated professionals with doctoral degrees representing 57% of respondents, likely contributes to this convergence. Education level and scientific training may act as equalising factors that override regional and demographic variation in technology acceptance.

### 4.4 Academic researchers versus healthcare professionals

A differential adoption pattern was identified between academic researchers and healthcare professionals, with the latter reporting lower hype perception and higher ML maturity in their field.

This divergence likely reflects structural differences between the two contexts. Academic researchers operate in environments where ML publications are abundant, and adoption is driven by scientific curiosity, while healthcare professionals face regulatory constraints, data quality challenges, and workflow integration barriers that create a more measured adoption trajectory (Da Silva et al., 2022; Pumplun et al., 2021).

This finding supports calls for subsector-specific ML adoption strategies. Interventions designed for academic researchers, emphasising cutting-edge applications and publication-driven evidence, may be ineffective in clinical settings, where regulatory compliance, interoperability with existing systems, and demonstrated patient outcomes are the primary adoption drivers.

Beyond individual acceptance variables, the broader implications of ML integration in life science organisations warrant further theoretical exploration. Emerging perspectives on algorithmic trust, human-AI collaboration, and epistemic legitimacy in scientific decision-making represent important dimensions not fully captured by the TAM framework alone. Future research should examine how institutional dynamics, professional identity, and governance structures interact with individual acceptance factors to shape organisational ML adoption trajectories, particularly in highly regulated environments where socio-technical transformation occurs incrementally and under significant compliance constraints.

## 5. Conclusions

This study investigated machine learning technology acceptance among biomedical and life science professionals using an extended TAM framework incorporating three external variables: journal impact factor, technology hype perception, and prior knowledge and experience. Based on a cross-sectional quantitative survey of 213 professionals across EMEA, LATAM, and North America, all ten hypotheses were supported, confirming both the core TAM structure and the positive influence of the three external variables on perceived usefulness and perceived ease of use.

The primary theoretical contribution is the validation of JIF and technology hype as industry-specific antecedents of ML acceptance in life sciences, a sector where bibliometric culture and hype cycle dynamics shape professional decision-making in ways not captured by generic TAM extensions. The finding that ML adoption in life sciences follows a different hype trajectory than general IT industries further underscores the importance of sector-specific technology acceptance research.

Practically, the findings suggest that organisations seeking to accelerate ML implementation should invest in building behavioural intention through hands-on training and peer exposure, disseminate ML evidence through high-impact scientific journals, and develop sector-differentiated adoption strategies that account for the distinct dynamics of academic research versus clinical environments.

### 5.1 Limitations and future research

Several limitations of this study should be acknowledged. First, the convenience sampling approach via LinkedIn introduces selection bias; respondents who are active on professional networks may hold more favourable attitudes toward technology adoption than the broader life science professional population. Second, the relatively modest sample size of 213 respondents, while sufficient for PLS-SEM analysis, limits the granularity of subsector-specific analyses. Third, the cross-sectional design precludes causal inference; the relationships identified are associative rather than directional over time. Fourth, the weak R-squared values for ATU (0.197) and PEU (0.159) suggest that additional variables beyond those included in the model may explain variance in these constructs. Fifth, while the TAM framework provides a well-validated foundation for technology acceptance research, future studies should consider complementary theoretical lenses, including socio-technical systems theory, institutional theory, and human-AI collaboration frameworks, to capture the broader organisational and governance dimensions of ML adoption in life sciences.

Future research should address these limitations through larger, probability-based samples with greater subsector granularity, particularly separating clinical diagnostics, pharmaceutical R&D, and academic research into distinct study populations. Longitudinal designs would allow examination of how ML acceptance evolves as the technology matures within the sector. Additionally, the post-2022 emergence of large language models and generative AI has substantially altered the ML landscape in life sciences. Replication of this study with updated instruments would provide valuable comparative data on how the rapid proliferation of accessible AI tools has shifted adoption perceptions among biomedical professionals

## Ethics statement

This study was conducted in accordance with the ethical guidelines of the University of Winchester (Research Ethics Form 3, approved December 2022, supervisor: Tim Friesner, Faculty of Business and Digital Technologies). All participation was voluntary and anonymous. No personally identifiable information was collected. Informed consent was obtained from all participants prior to data collection. The study was classified as low risk and did not require additional institutional review board approval beyond the university ethics process.

## Data availability statement

The anonymised dataset supporting the findings of this study is available on Harvard Dataverse: https://doi.org/10.7910/DVN/F3BPVF

## Declaration of generative AI use

During the preparation of this manuscript, the author used Claude (Anthropic, claude.ai) to assist with manuscript restructuring from dissertation to journal article format, including abstract reformatting and section condensation. The author reviewed and edited all AI-assisted content and takes full responsibility for the content of the submitted article.

## CRediT author statement

Antonio E. Serrano: Conceptualization, Data curation, Formal analysis, Investigation, Methodology, Project administration, Visualization, Writing original draft, Writing review and editing.

---

**Supplementary Material 1.**
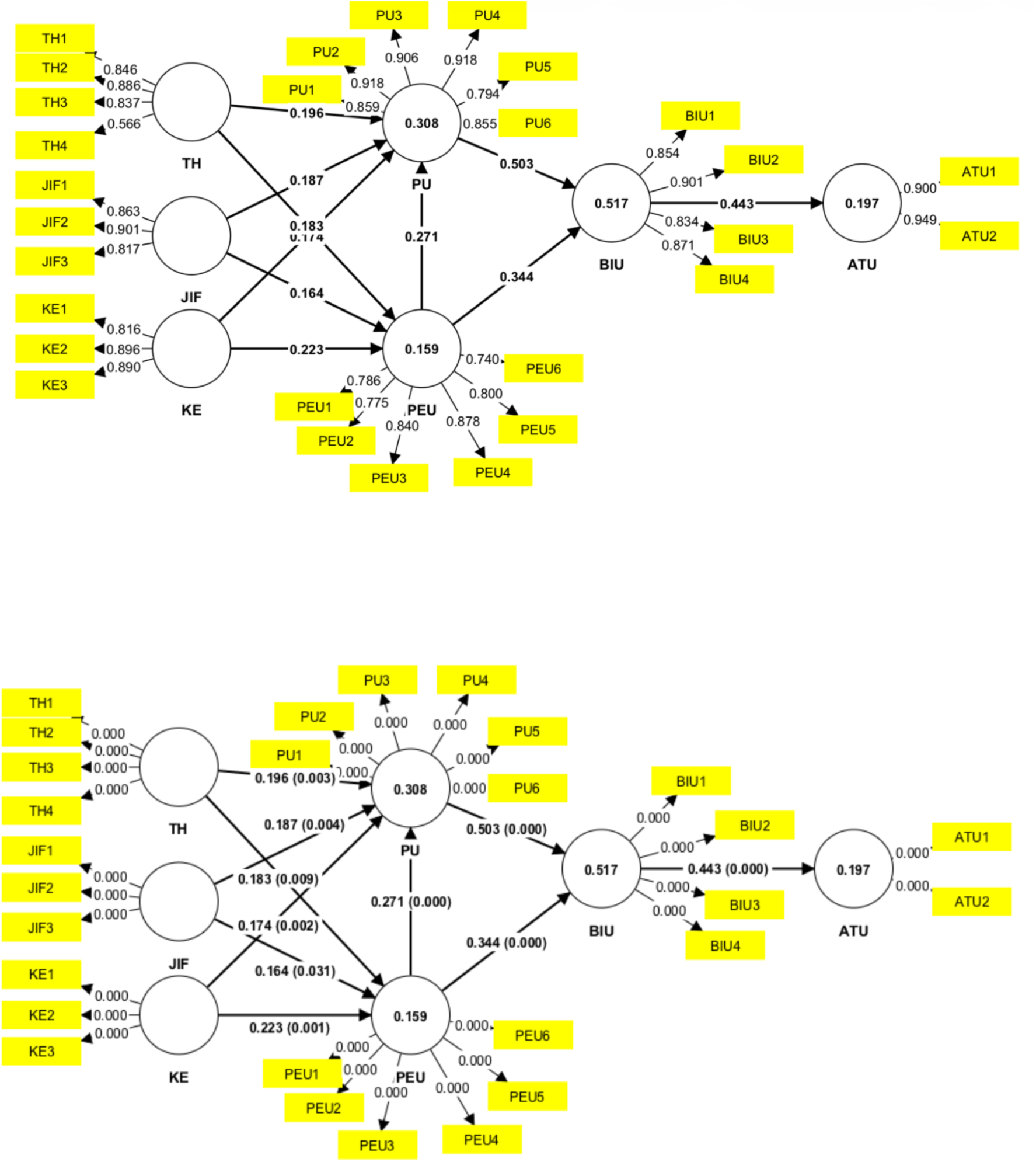
PLS-SEM graphic models.

**Supplementary Material 2.**
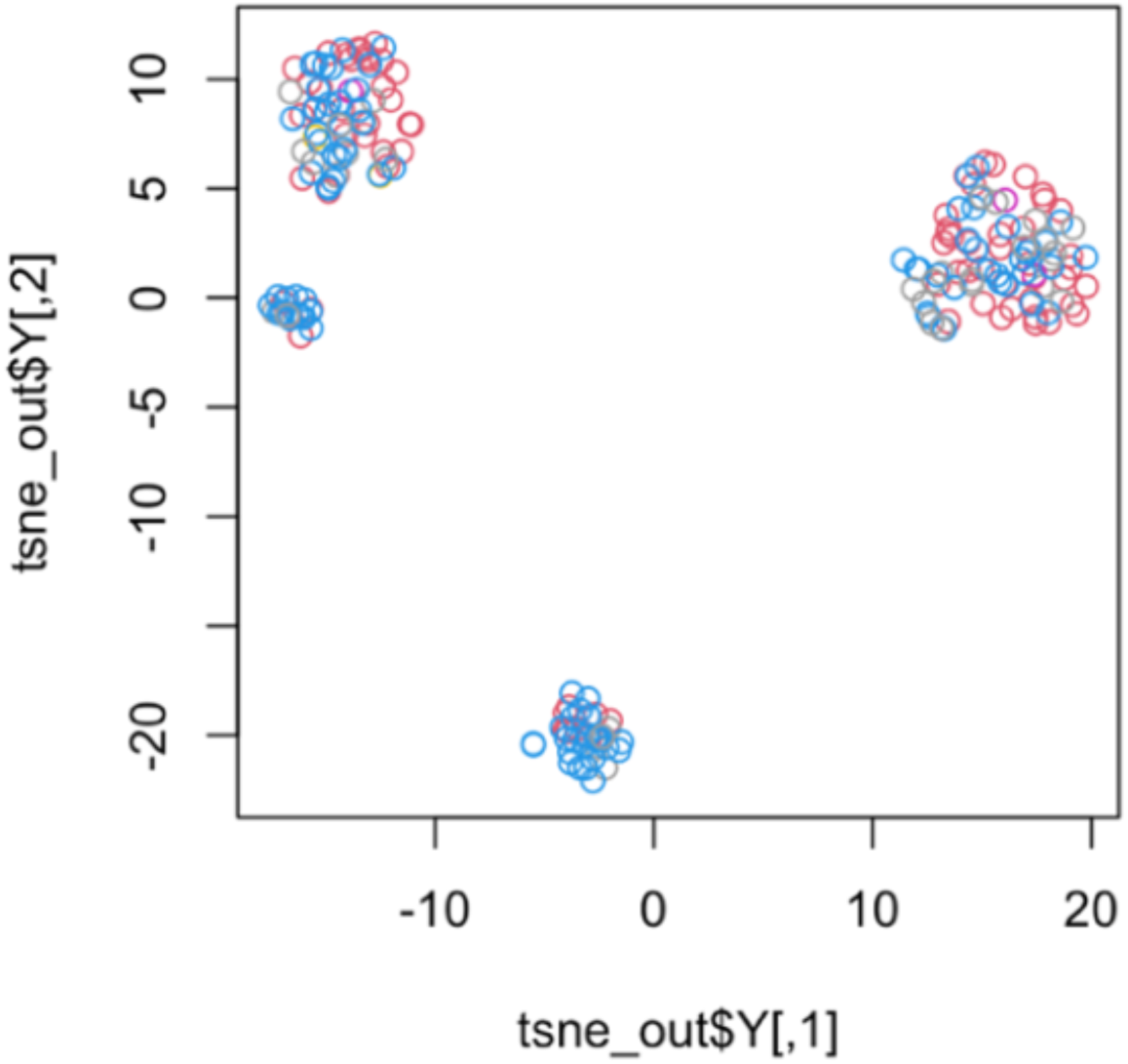
t-SNE test analysis.

**Supplementary Material 3.**
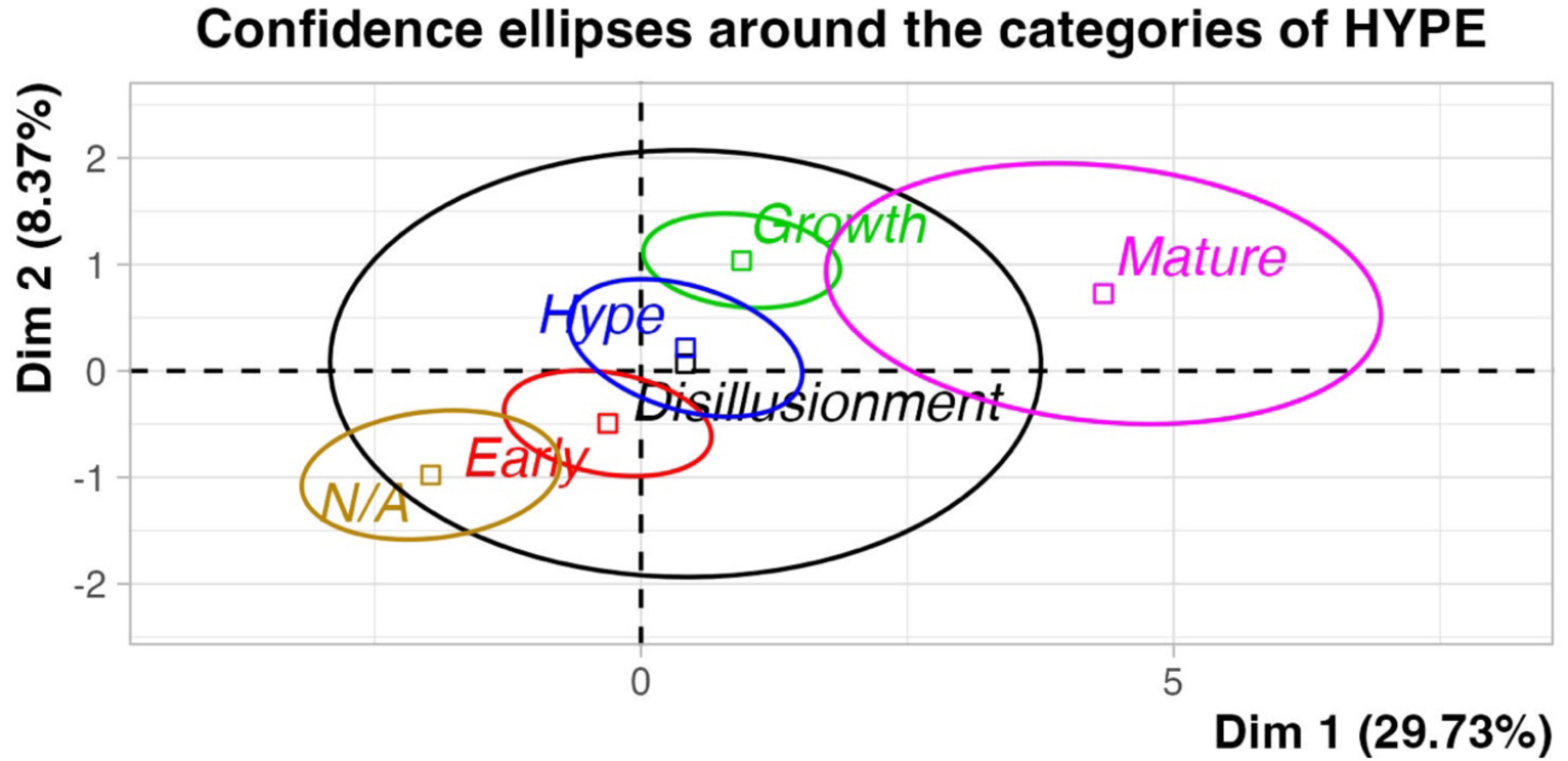
MCA analysis. Confidence ellipses of the hype plot.

## References

Alalwan, A. A., Dwivedi, Y. K., Rana, N. P., & Algharabat, R. (2018). Examining adoption of mobile internet in Saudi Arabia: Extending TAM with perceived enjoyment, innovativeness and trust. Technology in Society, 55, 100–110. 10.1016/j.techsoc.2018.06.007

Andrews, J. E., Ward, H., & Yoon, J. W. (2021). UTAUT as a model for understanding intention to adopt AI and related technologies among librarians. Journal of Academic Librarianship, 47(3), Article 102437. 10.1016/j.acalib.2021.102437

Baltimore, D. (2001). How biology became an information science. In M. Dertouzos (Ed.), The invisible future: The seamless integration of technology into everyday life (pp. 43–55). McGraw Hill.

Balyen, L., & Peto, T. (2019). Promising artificial intelligence–machine learning–deep learning algorithms in ophthalmology. Asia-Pacific Journal of Ophthalmology, 8(3), 263–271. 10.22608/APO.2018479

Benchaaben, A., Guimaraes, F. M., Prestat, E., Kassambara, A., Filahi, M., & Laugé, C. (2020). Abstract 870: Immunoscore® workflow enhanced by artificial intelligence. Cancer Research, 80(16). 10.1158/1538-7445.am2020-870

Bresciani, S., & Eppler, M. J. (2010). Gartner’s magic quadrant and hype cycle. Collaborative Knowledge Visualization Case Study Series.

Bzdok, D., Altman, N., & Krzywinski, M. (2018). Statistics versus machine learning. Nature Methods, 15(4), 233–234. 10.1038/nmeth.4642

Chen, M., Zhang, B., Cai, Z., Seery, S., Mendez, M. J., Ali, N. M., Ren, R., Qiao, Y. L., Xue, P., & Jiang, Y. (2022). *Physician and medical student attitudes toward clinical artificial intelligence: A systematic review with cross-sectional survey* [Preprint]. SSRN. 10.2139/ssrn.4128867

Da Silva, M., Camargo, L., Rodrigues, R., Silva, A., Mendes, R., & Gonçalves, R. (2022). Legal concerns in health-related artificial intelligence: A scoping review protocol. Systematic Reviews, 11(1), Article 123. 10.1186/s13643-022-01939-y

Davenport, T., & Kalakota, R. (2019). The potential for artificial intelligence in healthcare. Future Healthcare Journal, 6(2), 94–98. 10.7861/futurehosp.6-2-94

Davis, F. D. (1985). A technology acceptance model for empirically testing new end-user information systems: Theory and results [Doctoral dissertation]. MIT Sloan School of Management.

Davis, F. D. (1989). Perceived usefulness, perceived ease of use, and user acceptance of information technology. MIS Quarterly, 13(3), 319–340. 10.2307/249008

Dey, A. (2016). Machine learning algorithms: A review. International Journal of Computer Science and Information Technologies, 7(3), 1174–1179.

Fiala, J., Mareš, J. J., & Šesták, J. (2017). Reflections on how to evaluate the professional value of scientific papers and their corresponding citations. Scientometrics, 112(1), 697–709. 10.1007/s11192-017-2334-x

Florensa, D., Aguilera, P., Roca, M., & Meléndez, M. (2021). The use of multiple correspondence analysis to explore associations between categories of qualitative variables and cancer incidence. IEEE Journal of Biomedical and Health Informatics, 25(6), 2015–2025. 10.1109/JBHI.2021.3073605

Fornell, C., & Larcker, D. F. (1981). Evaluating structural equation models with unobservable variables and measurement error. Journal of Marketing Research, 18(1), 39–50. 10.1177/002224378101800104

Garfield, E. (1996). How can impact factors be improved? BMJ, 313(7054), 411–413. 10.1136/bmj.313.7054.411

Gartner Inc. (2018). Gartner hype cycle: Interpreting technology hype. Gartner.

Goasduff, L. (2020). 2 megatrends dominate the Gartner hype cycle for artificial intelligence, 2020. Gartner. https://www.gartner.com/en/articles/2-megatrends-dominate-the-gartner-hype-cycle-for-artificial-intelligence-2020

Goasduff, L. (2021). The 4 trends that prevail on the Gartner hype cycle for AI, 2021. Gartner. https://www.gartner.com/en/articles/the-4-trends-that-prevail-on-the-gartner-hype-cycle-for-ai-2021

Goecks, J., Jalili, V., Heiser, L. M., & Gray, J. W. (2020). How machine learning will transform biomedicine. Cell, 181(1), 92–101. 10.1016/j.cell.2020.03.022

Gong, L. (2018). Application of biomedical text mining. In S. Kountchev & K. Nakamatsu (Eds.), Artificial intelligence: Emerging trends and applications. IntechOpen. 10.5772/intechopen.75924

Gu, J., Xu, Y. C., Xu, H., Zhang, C., & Ling, H. (2015). Construction of a technology adoption decision-making model and its extension to understanding herd behavior. Knowledge-Based Systems, 85, 111–124. 10.1016/j.knosys.2015.08.014

Hair, J. F., Risher, J. J., Sarstedt, M., & Ringle, C. M. (2019). When to use and how to report the results of PLS-SEM. European Business Review, 31(1), 2–24. 10.1108/EBR-11-2018-0203

Harrison, J. H., Gilbertson, J. R., Hanna, M. G., Olson, N. H., Seheult, J. N., Sorace, J. M., & Parwani, A. V. (2021). Introduction to artificial intelligence and machine learning for pathology. Archives of Pathology and Laboratory Medicine, 145(10), 1228–1254. 10.5858/arpa.2020-0541-CP

Henseler, J., Ringle, C. M., & Sarstedt, M. (2015). A new criterion for assessing discriminant validity in variance-based structural equation modeling. Journal of the Academy of Marketing Science, 43(1), 115–135. 10.1007/s11747-014-0403-8

Hong, S. H., & Yu, J. H. (2018). Identification of external variables for the Technology Acceptance Model (TAM) in the assessment of BIM application for mobile devices. IOP Conference Series: Materials Science and Engineering, 401, Article 012027. 10.1088/1757-899X/401/1/012027

Horowitz, M. C., & Kahn, L. (2021). What influences attitudes about artificial intelligence adoption: Evidence from U.S. local officials. PLOS ONE, 16(2), Article e0257732. 10.1371/journal.pone.0257732

Huang, K., Bhatt, D. L., Bhattacharya, S., & Leiter, R. E. (2021). Machine learning applications for therapeutic tasks with genomics data. Patterns, 2(8), Article 100328. 10.1016/j.patter.2021.100328

Hubona, G. S., & Whisenand, T. G. (1995). External variables and the Technology Acceptance Model. Georgia State University.

Hultberg, J., Lonnroth, K., & Makinen, M. (1999). An international comparison of technology adoption and efficiency: A dynamic panel model. Annales d’Économie et de Statistique, 55/56, 389–414. 10.2307/20076207

Jimenez, I. A. C., García, L. C. C., Violante, M. G., Marcolin, F., & Vezzetti, E. (2021). Commonly used external TAM variables in e-learning, agriculture and virtual reality applications. Future Internet, 13(1), Article 7. 10.3390/fi13010007

Jin, M. C., Krishnan, V., & Prolo, L. (2022). A discussion of machine learning approaches for clinical prediction modeling. In *Acta Neurochirurgica*, Supplementum (Vol. 134, pp. 57–63). Springer. 10.1007/978-3-030-85292-4_9

Kannan, R., Garg, N., Gupta, A., & Bhardwaj, A. (2021). Leveraging business data analytics and machine learning techniques for competitive advantage. *International Journal of Management*, Finance and Accounting, 2(1), 1–15. 10.33093/ijomfa.2021.2.1.3

Kaur, A., & Malik, G. (2019). Examining factors influencing Indian customers’ intentions and adoption of internet banking: Extending TAM with electronic service quality. Innovative Marketing, 15(2), 42–57. 10.21511/im.15(2).2019.04

Koohy, H. (2018). The rise and fall of machine learning methods in biomedical research. F1000Research, 7, Article 1213. 10.12688/f1000research.13016.2

Lê, S., Josse, J., & Husson, F. (2008). FactoMineR: An R package for multivariate analysis. Journal of Statistical Software, 25(1), 1–18. 10.18637/jss.v025.i01

Liu, G., Zeng, H., Mueller, J., Carter, B., Wang, Z., Schilter, J., & Rampasek, L. (2020). Antibody complementarity determining region design using high-capacity machine learning. Bioinformatics, 36(7), 2126–2133. 10.1093/bioinformatics/btz895

Nurk, S., Koren, S., Rhie, A., Rautiainen, M., Bzikadze, A. V., Mikheenko, A., & Phillippy, A. M. (2021). The complete sequence of a human genome. bioRxiv. 10.1101/2021.05.26.445798

Patel, L., Shukla, T., Huang, X., Ussery, D. W., & Wang, S. (2020). Machine learning methods in drug discovery. Molecules, 25(22), Article 5277. 10.3390/molecules25225277

Phung, D., Webb, G. I., & Sammut, C. (Eds.). (2020). Encyclopedia of machine learning and data science. Springer. 10.1007/978-1-4899-7502-7

Posit Team. (2022). RStudio: Integrated development for R (Version 2022.12.0.353) [Computer software]. Posit Software. http://www.posit.co/

Pumplun, L., Fecho, M., Wahl, N., Peters, F., & Buxmann, P. (2021). Adoption of machine learning systems for medical diagnostics in clinics: Qualitative interview study. Journal of Medical Internet Research, 23(10), Article e29301. 10.2196/29301

Rajkumar, D. (2020). Applications of machine learning in radiology: A review. Journal for Innovative Development in Pharmaceutical and Technical Science, 3(6), 1–8.

Rashidi, H. H., Tran, N. K., Betts, E. V., Howell, L. P., & Green, R. (2019). Artificial intelligence and machine learning in pathology: The present landscape of supervised methods. Academic Pathology, 6, 1–11. 10.1177/2374289519873088

Ringle, C. M., Wende, S., & Becker, J. M. (2022). SmartPLS 4 [Computer software]. SmartPLS. https://www.smartpls.com

Rogers, E. M. (1962). Diffusion of innovations. Free Press of Glencoe.

Sarker, I. H. (2021). Machine learning: Algorithms, real-world applications and research directions. SN Computer Science, 2(3), Article 160. 10.1007/s42979-021-00592-x

Saunders, M., Lewis, P., & Thornhill, A. (2009). Research methods for business students (5th ed.). Pearson Education.

Shi, K., Lin, W., & Zhao, X. M. (2021). Identifying molecular biomarkers for diseases with machine learning based on integrative omics. IEEE/ACM Transactions on Computational Biology and Bioinformatics, 18(2), 440–450. 10.1109/TCBB.2020.2986387

Sodhi, M. M. S., Tang, C. S., & Willenson, E. T. (2022). Why emerging supply chain technologies initially disappoint: Blockchain, IoT, and AI. Production and Operations Management, 32(4), 1163–1180. 10.1111/poms.13694

Van der Maaten, L. J. P., & Hinton, G. E. (2008). Visualizing high-dimensional data using t-SNE. Journal of Machine Learning Research, 9, 2579–2605.

Venkatesh, V., & Davis, F. D. (1996). A model of the antecedents of perceived ease of use: Development and test. Decision Sciences, 27(3), 451–481. 10.1111/j.1540-5915.1996.tb00860.x

Venkatesh, V., & Davis, F. D. (2000). Theoretical extension of the Technology Acceptance Model: Four longitudinal field studies. Management Science, 46(2), 186–204. 10.1287/mnsc.46.2.186.11926

Vincent, J. (2019, March 5). Forty percent of ‘AI startups’ in Europe don’t actually use AI, claims report. The Verge. https://www.theverge.com/2019/3/5/18251326/ai-startups-europe-fake-40-percent-mmc-report

Watkins, M. W. (2018). Exploratory factor analysis: A guide to best practice. Journal of Black Psychology, 44(3), 219–246. 10.1177/0095798418771807

World Bank. (2022). GNI per capita, PPP (current international $). World Bank International Comparison Program. https://data.worldbank.org/indicator/NY.GNP.PCAP.PP.CD

Xu, C., & Jackson, S. A. (2019). Machine learning and complex biological data. Genome Biology, 20(1), Article 76. 10.1186/s13059-019-1689-0

Zhang, X., Wang, Y., & Li, Z. (2021). User acceptance of machine learning models: Integrating several important external variables with technology acceptance model. International Journal of Electrical Engineering Education. 10.1177/00207209211005271

